# Examination of Isolation Rate in SIQR model for COVID-19 Epidemic

**DOI:** 10.1101/2020.09.01.20185611

**Authors:** Koichi Hashiguchi

## Abstract

Newly proposed SIQR model defines exponent *λ* of exponential function expressing daily number of isolated persons as linear equation of isolation rate q and social distancing ratio x. In order to dynamically analyze the process of COVID-19 epidemic in seven countries by means of regression analyses of *λ*, increasing rate of cumulative isolated persons(cases), IRCC, is proposed as practical index for the isolation rate q. IRCC is correlated with q in the form of q=C · IRCC, where C is a normalizing coefficient. At first, C is formulated in two modes, one is simple and the other complex, under the constraint conditions by definition 0≤x, q≤1, which give allowable narrow path of C between upper and lower boundaries. Then, the dynamic locus of q-x relation is analyzed for each of seven countries including Japan and the United States using formulated isolation rate q, and characteristic q-x behavior for each country is derived. At the same time, it is shown that specific path selection of C gives almost same linear loci of q-x relation derived by mathematical sequential method imitating a bipedal walk. In addition, increasing rates of cumulative PCR tests, IRCT, for six countries are discussed in relation with IRCC, and are shown that IRCT contributes to the promotion of the isolation rate via IRCC.

## Introduction

The applicability of newly proposed SIQR model by Odagaki^1^ was verified by dynamic analyses^2^) using publicly available database for typical seven countries. *λ*, the exponent of exponential function for daily new cases, is defined as a linear expression of isolation rate q and social distancing ratio x, which is the basic equation in SIQR model and indefinite. This indefinite equation was solved mathematically by sequential method imitating so-called bipedal walk without any practical and physical consideration for the isolation rate^2)^. As a result, the locus of q-x relation during the progress of infection for each country was able to be determined.

The isolation rate must be a practical and physical meaningful parameter from a viewpoint of practical use. Therefore, the increasing rate of cumulative cases (IRCC/day) is proposed as an index with a practical meaning as the isolation rate. First in this work, the analyzing method and database used in the previous report are briefly reviewed. Then normalizing coefficient C is introduced to correlate isolation rate q and IRCC as q·CIRCC. Further, C is formulated in two modes, one is simple and the other complex, under the constraint conditions by definition of 0≤x, q≤1, which give allowable narrow path of C during infection period between upper and lower boundaries of C. Then, the dynamic locus of q-x relation is analyzed for each of seven countries including Japan and the United States using formulated isolation rate q, and characteristic q-x behavior of each country is derived. At the same time, it is shown that specific path selection of C gives almost same loci of linear q-x relation derived mathematically in the previous report. In addition, increasing rates of cumulative PCR tests, IRCT, for six countries are discussed in relation with IRCC, and are shown that IRCT contributes to the promotion of the isolation rate via IRCC.

### Basic equation in SIQR model and approaching method in data-fitting

The same analyzing method as previous analyses was taken, the important items of which are reviewed briefly in the following.

- Basic equation; The number of daily isolated persons (tested as positive and maybe isolated, hereafter new cases) ΔQ is expressed by logarithmic approximation formula (1) with constant term *ΔQ*_0_, and λ value was obtained as a coefficient of linear regression. The regression analyzed daily *λ* leads to the dynamic capturing of transition of infection process.

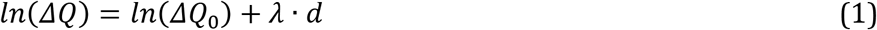
- Definition of exponent λ; In the SIQR model, the exponent λ of eq. (1) is defined by eq. (2).

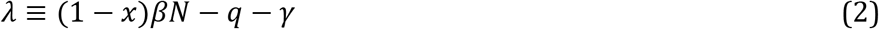 Here, βN is a value obtained by multiplying the transmission coefficient β by population N, an index of the degree of transmission through contact between uninfected and infected persons, x is a social distancing ratio, and q is an isolation rate. *γ* is the cure rate which was set equal to 0.04. The unit of *λ* is (1/day), q and βN have the same unit, and x is dimensionless.
- Approximation in the model; In each of the four groups of S(Susceptible), I(Infected), Q(Quarantined), and R(Recovered), the degree of impact on infection may differ due to depopulation, regional differences such as age composition, and differences in measures, even though, only the average behavior in each country was considered similarly with Odagaki’s method. Additionally, the epidemic process is regarded as same even though the possibility of genetically different viruses.
- Analytical procedure; Both the social distancing ratio x and the isolation rate q in eq. (2) are parameters that take values of 0 to 1 based on their definitions. When both are 0, that is, without any measures to suppress infection, the right side of eq. (2) becomes βN − γ, which gives the maximum value λ_max_ (= βN−γ), and the number of new cases increases at the fastest speed. This relationship is substituted into eq. (2) and transformed to obtain eq. (3).

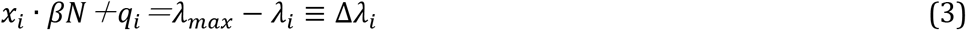 βN associated with the maximum λ is considered as constant throughout the infection period. Using the right-hand side Δλ of eq. (3) determined by *λ_max_* and daily λ, it is possible to determine the social distancing ratio x and the isolation rate q.
- Range of regression analyses; The regression analyses for daily λ, increasing rate of cumulative cases and increasing rate of cumulative tests are performed in the range of *[d_i_ − Δd, d_i_ + Δd*] at day *d_i_*, which are set to 19 days with Δd=9. The analysis period is from the day of λmax (start day) to the convergence day.
- Terminology; The terms frequently used throughout this paper are unified as follows. Newly infected and isolated persons as new cases, maximum number of new cases as peak, and the day of minimum number of new cases after peak day as the convergence day.
- Database; The data for following 7 countries are analyzed using the daily isolated cases (new cases) and PCR tests in more than 200 countries/regions in the "Our World in Data" database ^3)^. The numbers of new cases and PCR tests were converted to per million people for comparison by country, and also were moving averaged with one week range. Japan with social distancing policy, Taiwan and South Korea that quickly settled due to swift response, Sweden for mass immunity, Italy, Germany, and the United States suffering from explosive mass outbreak.

### Increasing rate of cumulative cases from the viewpoint of isolation rate

In the previous analysis, the isolation rate q and the social distancing ratio x were derived as the mathematical solution of the indefinite equation^2^)’ and linear relation between q and x were obtained. There was no consideration of isolation rate from a practical and physical point of view in those analyses, therefore it is necessary to explore the physical basis of isolation rate based on the practical index for practical application of SIQR model.

The isolation rate q defined in eq. (2) is the index representing the rate of isolation from the group of infected but unquarantined patients, which directly corresponds to the daily new cases. Considering two requirements for q, as of 0≤ q≤1 and the unit of (1/day), the increasing rate of daily new cases is not suitable because it can be positive or negative depending on the daily increase or decrease. Remaining candidate for isolation rate is the number of new cases per day, which is always 0 or more. However, this number fluctuates largely depending on the method of data tabulation even after a moving average. Therefore, the increasing rate of the cumulative cases (IRCC) calculated as a daily slope of the cumulative cases curve, is used as a candidate for isolation rate. IRCC is more suitable than the number of new cases per day for the purpose of capturing the overall trend with less fluctuation through a leveling effect by regression.

IRCC is regression analyzed as the daily slope of the cumulative cases curve, and is shown in Figs. 1a and b. The start, peak and convergence days are shown as dots in the figure, and the days at maximum increasing rate are located close to the peak days. IRCC is assumed to be proportional to the isolation rate q. Under this assumption, the isolation rate increases rapidly in all countries from the start day of maximum value λmax, when q is zero, and the maximum isolation rate is recorded near the peak day. Thereafter, the isolation rate decreases toward the convergence day.

**Fig. 1.**
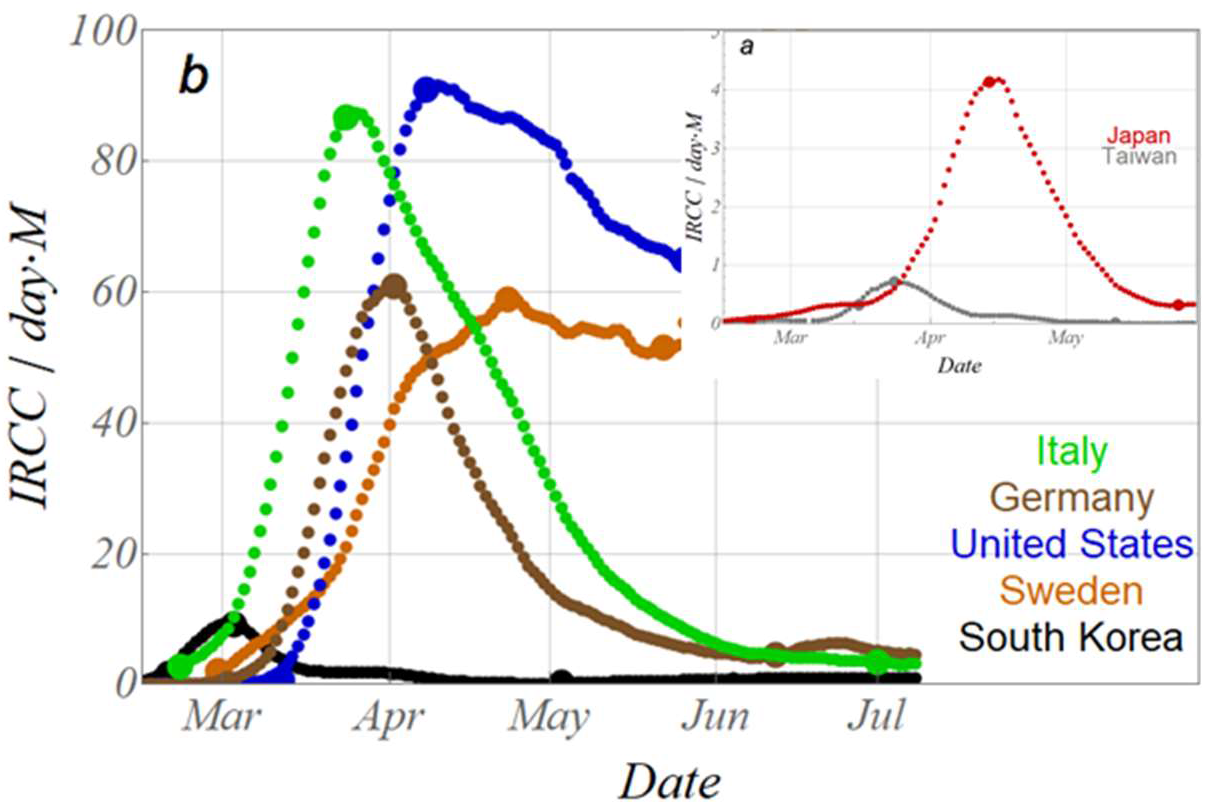
Variation of increasing rate of cumulative cases (IRCC) for seven countries, days at start, peak and convergence are marked with dots.

Let C be the coefficient that normalizes IRCC as the isolation rate q (0≤q≤1).

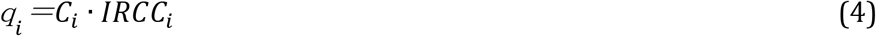

Substituting eq. (4) into eq. (3), and rearranging gives eq. (5) for the social distancing ratio xi.

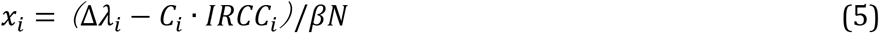

There is a constraint of 0≤ x_i_ ≤1, and eq. (5) is applied to this constraint to obtain eq. (6) for normalization of x_i_.

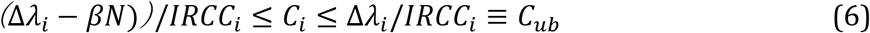

C_ub_ is defined as the upper boundary of C_i_. The upper and lower boundaries of C_i_ values are shown in Figs. 2a and 2b for Japan and Italy as typical examples of C_i_ values calculated using eq. (6). The normalization coefficient C_i_ is limited within these narrow upper and lower boundaries, and outside this range, the social distancing ratio becomes less than 0 or more than 1, which is not allowed by definition. Therefore, analyses are performed in two extreme cases in the range of C_i_ ≥ 0 (corresponding to q ≥ 0) after the start day. One is the case of the simple mode in which C_i_ value is set as 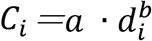 (*d_i_* is the number of days) so that it becomes a curve that passes through the neutral points between the upper and lower boundaries at around the peak day of new cases. The second case is a complex mode that traces upper boundary with 1/n value. C_i_ paths of these two cases are also shown in Figs. 2a and 2b.

**Fig. 2.**
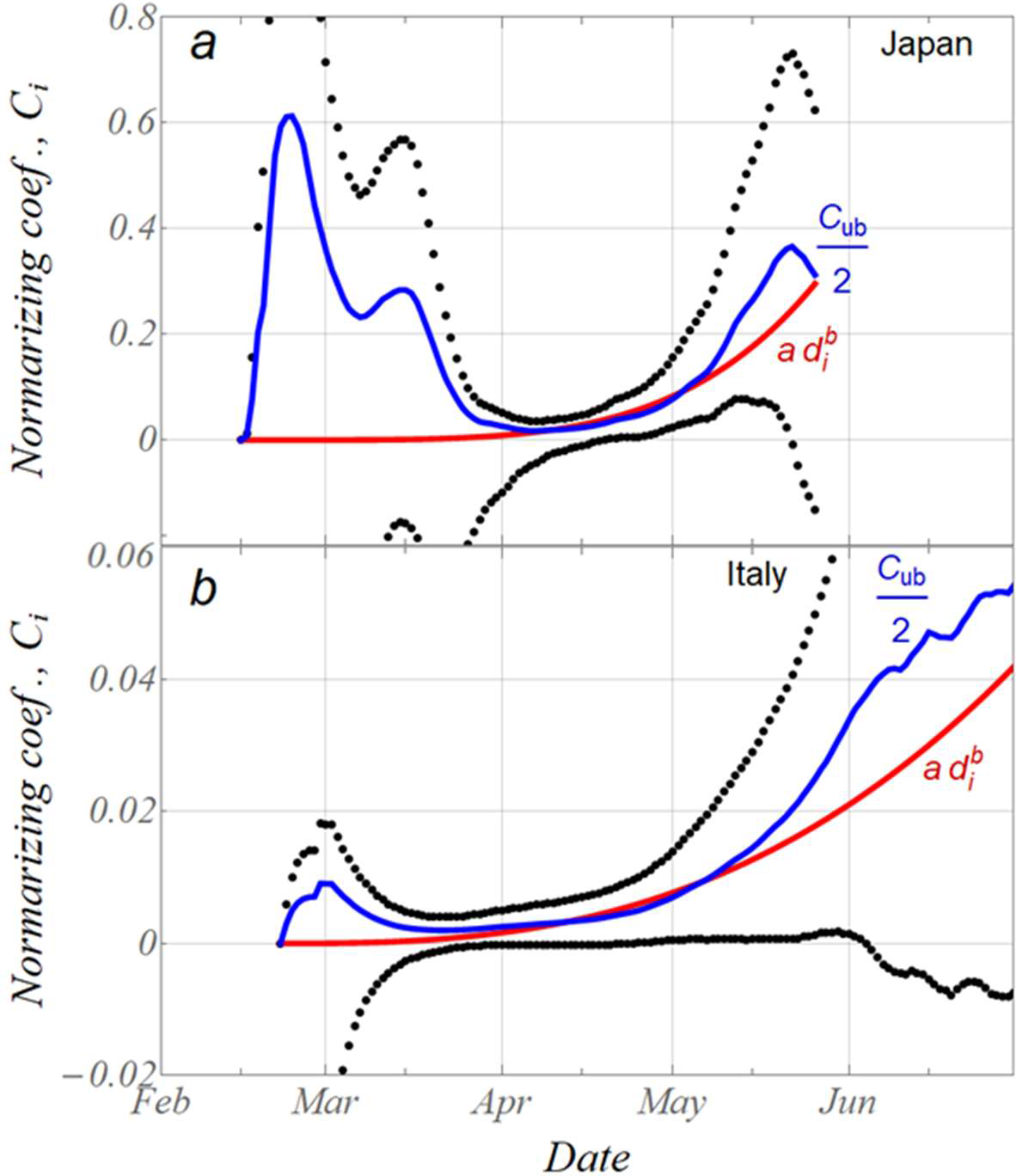
Upper and lower boundary curves of normalizing coef. *C_i_* for Japan (a) and Italy (b). *C_ub_ is* upper boundary curve for *C_i_*. Red and blue curves are for simple and complex mode, respectively.

**Fig. 3.**
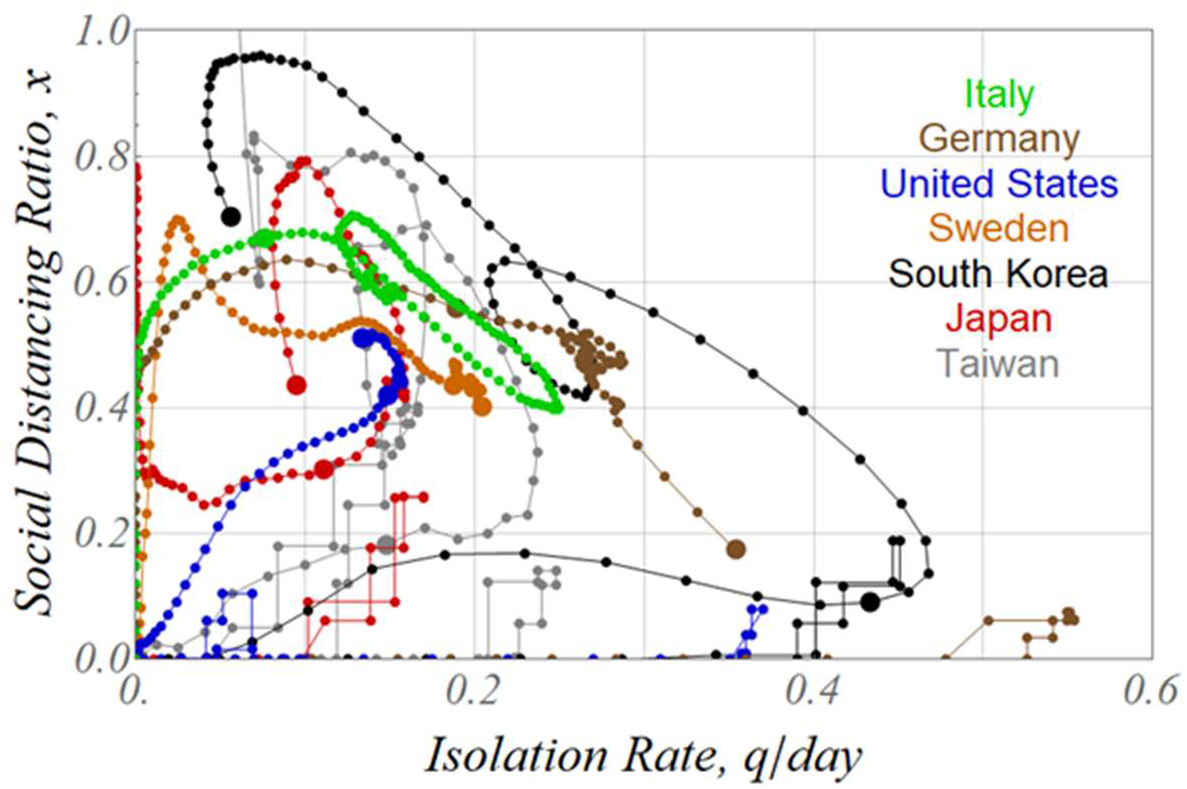
Moving locus of (q, x) point on q-x plane for seven countries, for simple mode with normalizing coef. *C_t_*, dots marked at peak and convergence days.

**Fig. 4.**
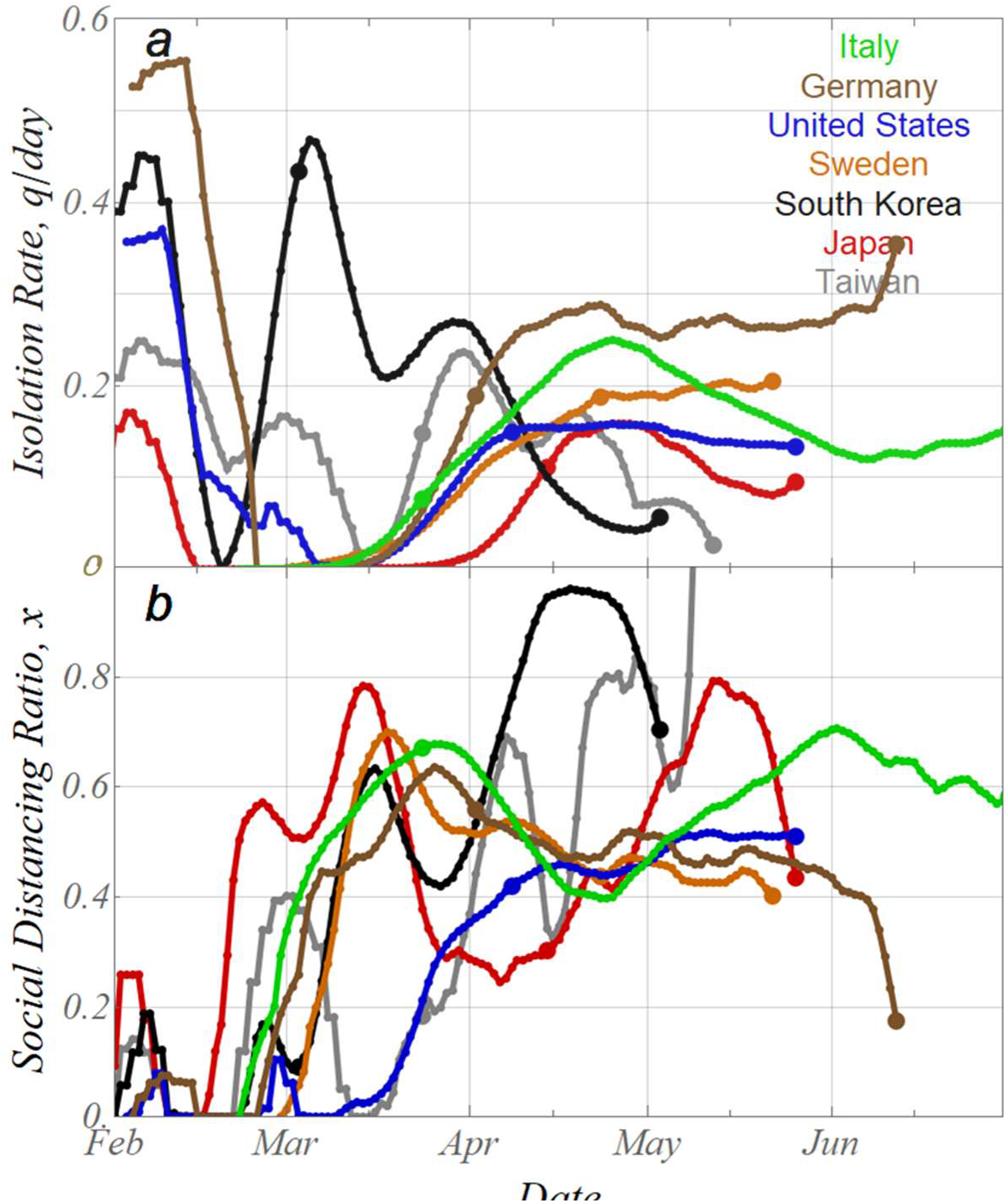
a, b Daily variation of isolation rate q (a) and social distancing ratio x (b) for seven countries.

In the case of the simple mode, the loci of each of the seven countries on the q-x coordinates calculated by eqs. (4) and (5) are shown in Fig. 3, and the daily variation of q and x for each country are shown in Figs. 4a and 4b. Variations of q and x at very early stages of infection even before λmax day calculated in the previous work are also shown in Figs. 3 and 4 as stepwise curves, which are not explained in this paper again and just remind the existence of sharp increase and decrease in isolation rate for some countries. Each locus on the q-x coordinate exhibits a complicated path. In Taiwan and South Korea, q increases significantly while x is low in the early stages of infection at the end of Feb., and q decreases while x increases after the peak. In Japan, x increases first, and then q increases. Germany, Sweden, and Italy exhibit similar variation with the Japanese type, however, only x increases with no initial increase in q, and q increases later. The US is characterized by the simultaneous increase in both of q and x and slight decrease even at the convergence point. Rapid increase of q in South Korea and also rapid increase of x in Italy and Germany at the end of Feb. may be caused by measures of each country’s policy such as intensive PCR test implementation in the former and lockdown in the latter. Assessment of these effects by various measures on q and x requires more detailed examination relating to time series of each countermeasures. In Taiwan, the social distancing ratio exceeds 1 in the middle of May (Fig. 4b), which corresponds to the decrease in C_i_ value crossing the lower boundary corresponding to this period. As a result of choice for the path of C_i_ as to pass through the neutral points between the upper and lower boundaries, the locus of q-x relation does not cross the upper and lower boundaries throughout the infection period. When the path of C_i_ deviate from the neutral point to the lower boundary side, the isolation rate decreases, and the social distancing ratio increases as shown in the case of Taiwan at middle May, and vice versa.

Figure 5 shows the locus on the q-x coordinate for the complex mode in which the C_i_ value is set to 1/2 of the upper boundary. In Fig. 5, unlike the complicated locus in Fig. 3, q and x follow very simple loci that change linearly, which is quite similar to the mathematical result calculated by the bipedal method in the previous report. In this complex mode, C_i_ is expressed by eq. (7) using 1/n instead of 1/2.

**Fig. 5.**
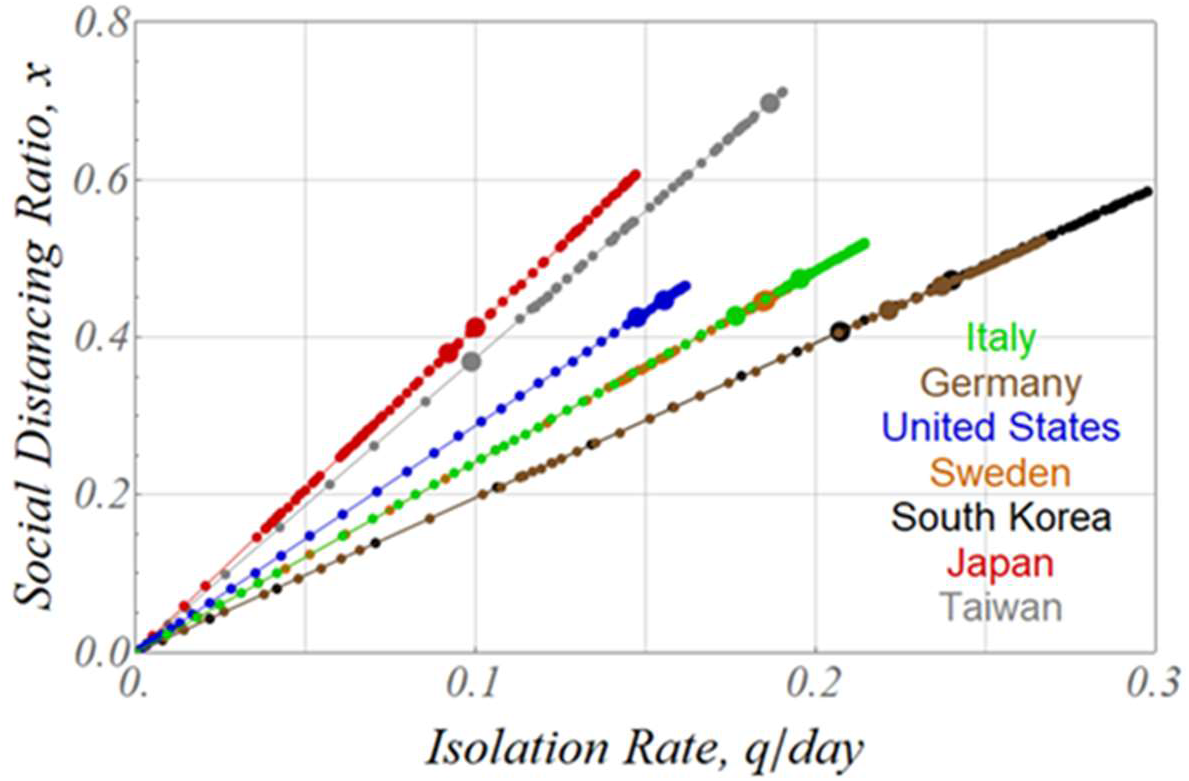
Moving locus of (q, x) point on q-x plane for seven countries, for complex mode with normalizing coef. *C_t_*, dots marked at peak and convergence days.

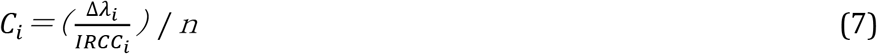

Substitution of eq. (7) into (4) gives *q_i_ =C_i_ · IRCC_i_ =Δλ_i_ / n*, which is substituted into eq. (3) or (5) to obtain eq. (8).

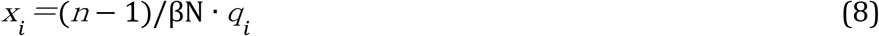

Therefore, the q-x relationship is a straight line of slope (n-1)/βN, and the differences of the slope in Fig. 5 are explained by the transmission coefficient βN for each country.

In this way, it became clear that the relationship between q and x varies greatly depending on the method of determining C_i_ value that links the isolation rate and IRCC. There is no guideline yet for determination of C_i_ path, which remains as a future analysis.

### Consideration of isolation rate from the viewpoint of number of PCR tests

The only means of controlling the isolation rate is to rely on tests such as PCR. The isolation rate is examined based on the viewpoint of PCR tests in this section. Although the definition of the number of PCR tests differs from country to country, such as the number of people tested or the number of tests, they were treated equally in this analysis.

Figure 6 shows the relationship between the number of daily new tests and the number of daily new cases for seven countries. In all countries, the number of new cases increases in proportion to the number of PCR tests during the increasing stage (small dots), and the number of new cases decreases with the number of tests remaining flat during the converging stage (a slightly different trend in the United States and Italy). This can be interpreted as follows.

**Fig. 6.**
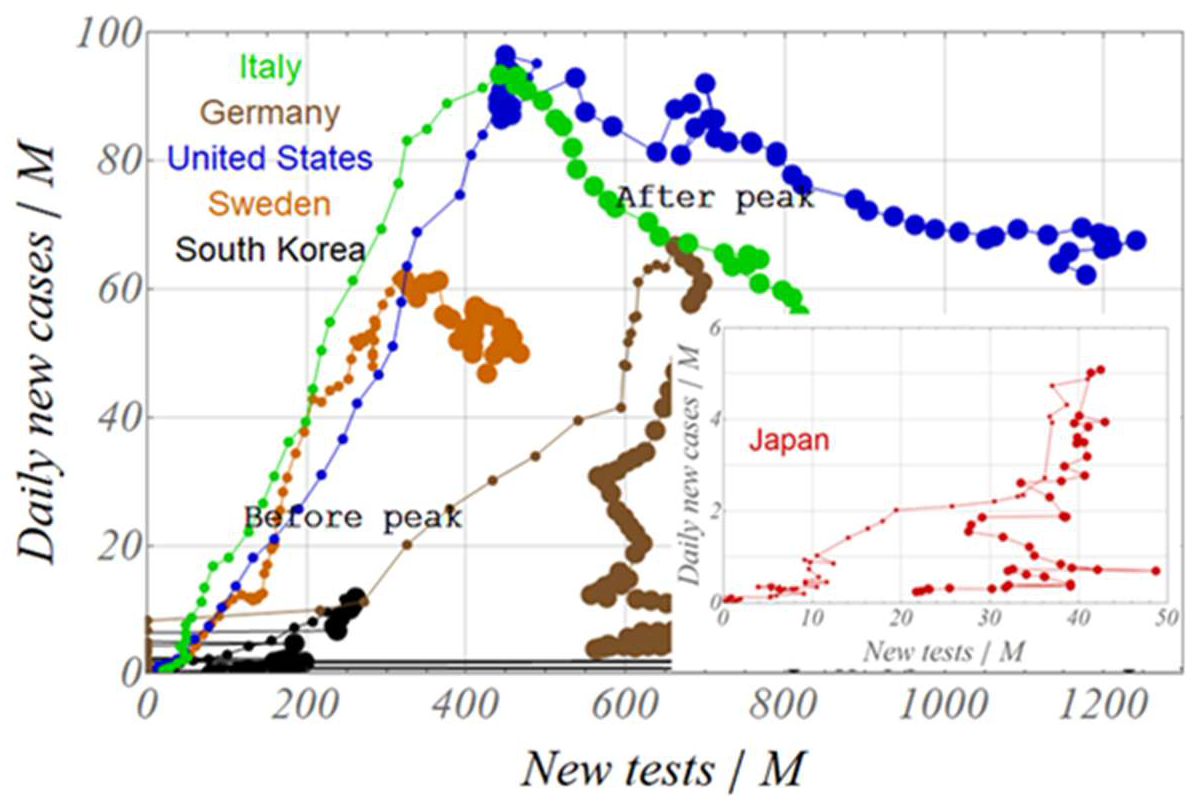
Relationship between daily new cases and daily new PCR tests for seven countries. Both numbers are converted to per million people for comparison by country.

The number of PCR tests increases in response to the increase in the number of new cases in each country during the increasing stage. Also increasing tests may increase the number of new cases. When the number of tests can catch up with the increasing number of new cases, then the number of new cases declines as the isolation progresses, making it unnecessary to increase the number of tests further. According to this interpretation, the tipping point for the increase/decrease in the number of new cases (which corresponds to the peak) can be regarded as the point at which the isolation rate becomes controllable. Therefore, this point can be considered as the saturation point of the isolation rate, as shown in Figs. 3 and 4 such that the peak days locate fairly close to the point of maximum q.

The relationship between the cumulative number of PCR tests and the cumulative number of new cases for seven countries is shown in Fig. 7. The dots are shown distinctively for the increasing stage (large dots) and the decreasing stage (small dots). The proportional relationship between the cumulative cases and the cumulative tests in the increasing stage continues up to the peak day, and then deviates from the proportional relation, indicating a tendency for the isolation to become saturated. Incidentally, the proportional constant between the two corresponds to the average positive ratio. Although the increase in the number of PCR tests leads to an increase in the number of isolation, the rate of increase in the number of tests on a daily basis is considered to be more important from the viewpoint of isolation control.

**Fig. 7.**
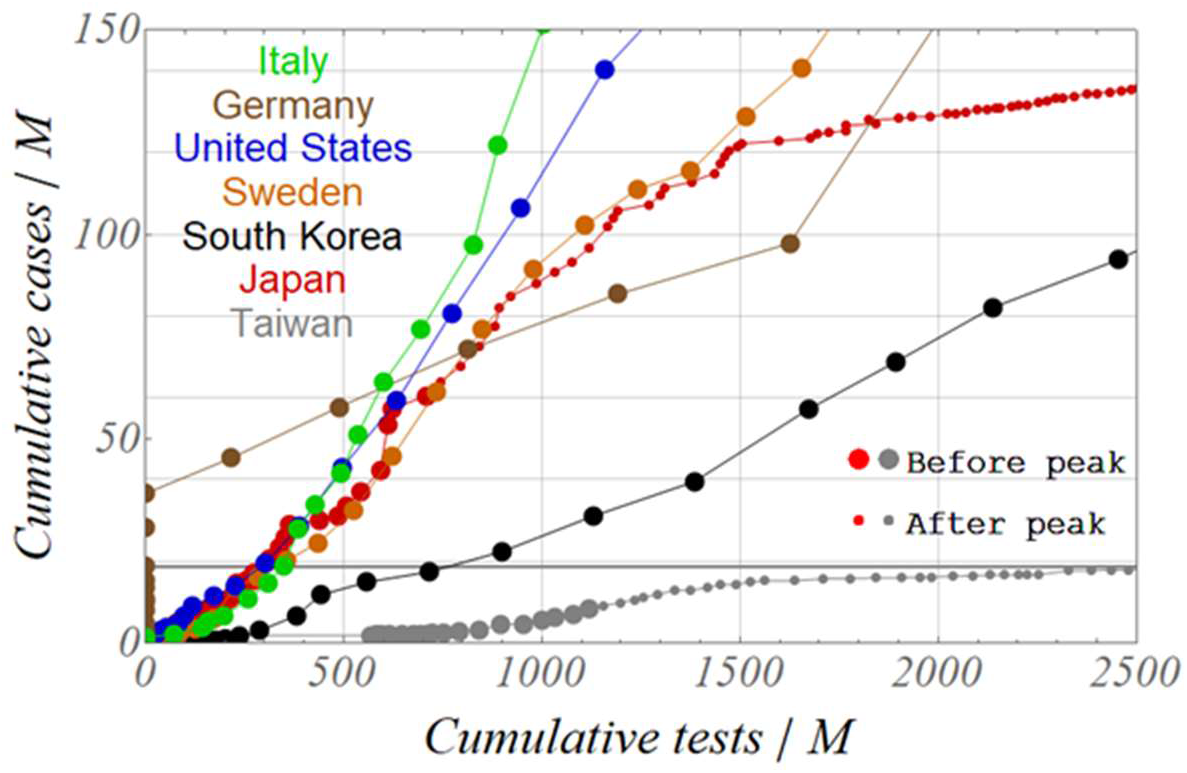
Relationship between cumulative PCR tests and cumulative cases for seven countries.

**Fig. 8.**
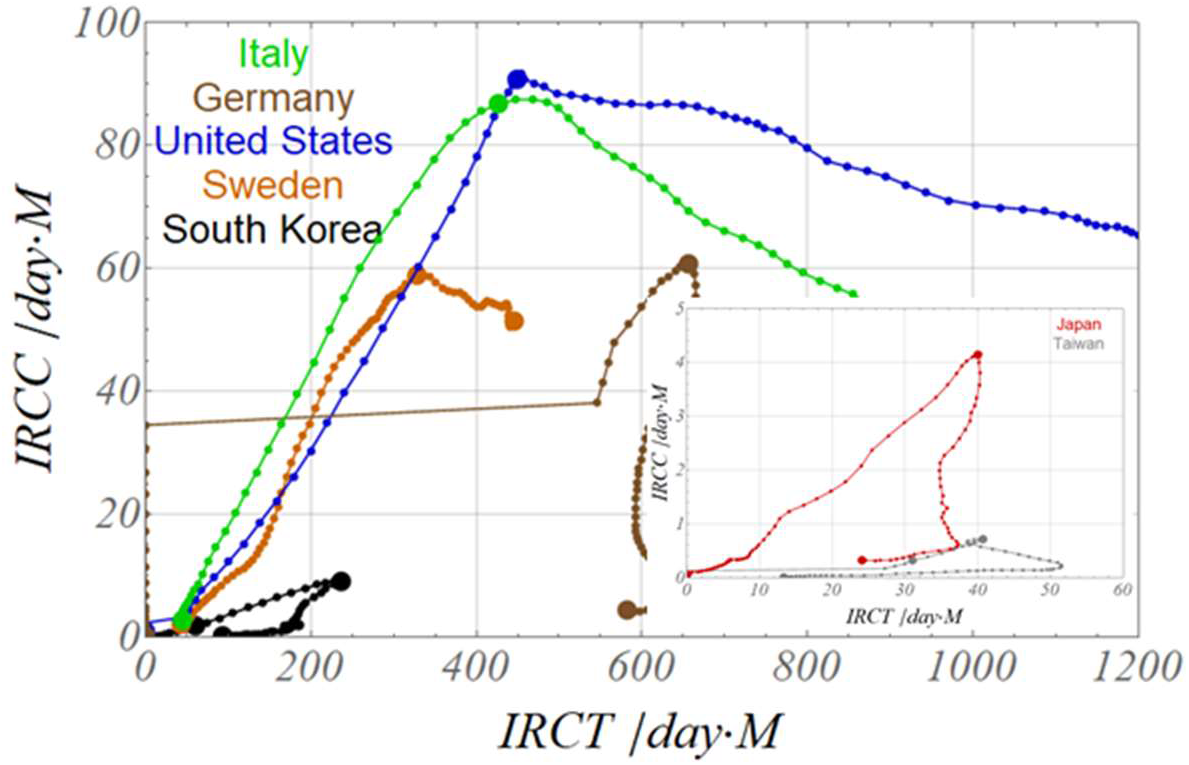
Relationship between increasing rate of cumulative cases (IRCC) and increasing rate of cumulative tests (IRCT) for seven countries, dots marked at peak days.

Figure 8 shows the relationship between IRCC and increasing rate of cumulative tests (IRCT) up to the convergence day. In each country, both of IRCC and IRCT increase linearly from the start of infection to the peak day, and IRCC, which corresponds to the isolation rate, decreases after the peak day. Germany was excluded from this analysis because of an unusual trend shown in the first half of the infection stage due to a problem in data tabulation. The slope of the linear relationship between IRCC and IRCT up to the peak is 0.04 for Taiwan and South Korea, and 0.1 for Japan, while the slope is a little more than 0.2 for Sweden, the United States and Italy. This slope is an indicator that expresses the ratio of the increase in the isolation rate per unit increase in IRCT, then can be regarded as a representative indicator of the isolation efficiency by the test. In fact, the relationship between this index and the average positive ratio of the proportional constant described in Fig. 7, are almost linear and show a positive correlation, as shown in Fig. 9. It can be concluded that the PCR tests were carried out efficiently in Europe and the United States where the average positive ratio is high. In Taiwan, Japan and South Korea with lower positive ratio, epidemic infection was controlled at low level even though the test efficiency is low, due to the low transmission coefficient in the former two countries as described in the previous report, and also due to the intensive test implementation at the early stage of infection in South Korea (Fig. 4b).

**Fig. 9.**
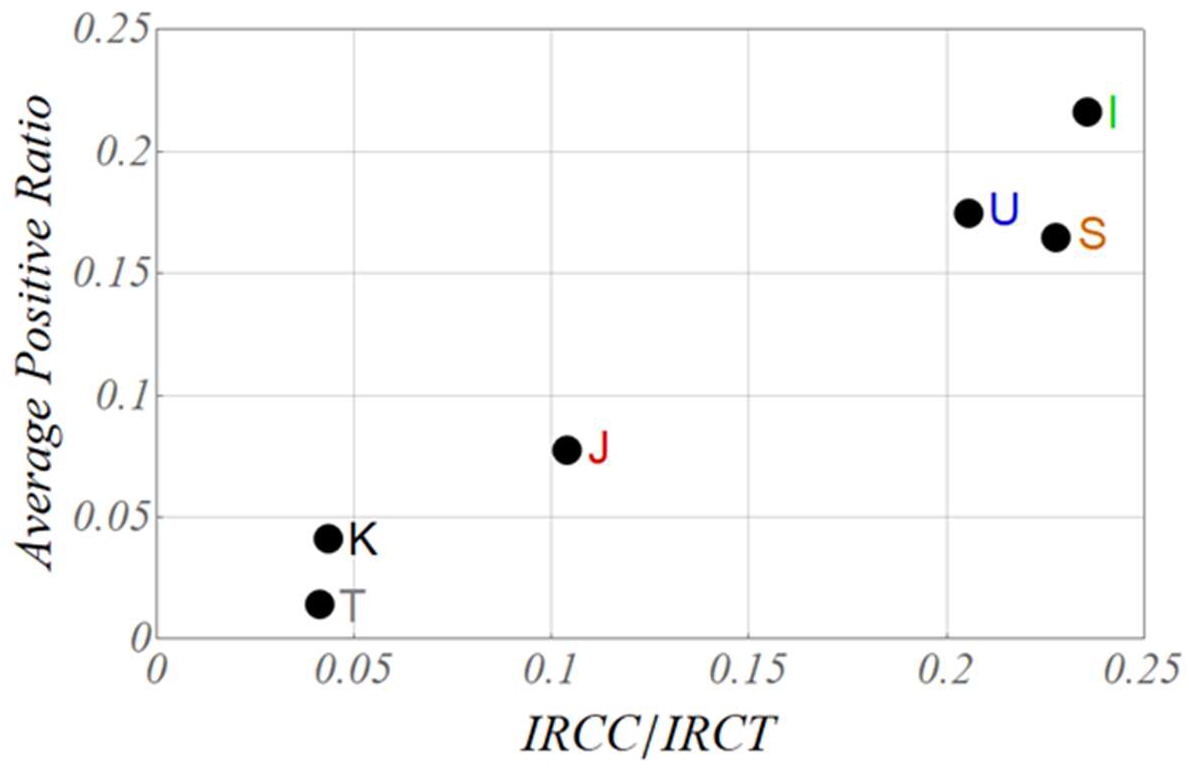
Relationship between ratio of increasing rate of cumulative cases (IRCC) to increasing rate of cumulative tests (IRCT) and average positive ratio for six countries, Taiwan, Korea, Japan, Sweden, USA and Italy.

## Conclusions

Throughout the previous report and this analysis, the process of COVID-19 epidemic in the seven countries was dynamically analyzed by applying newly proposed SIQR model to publicly available database through the analyses of exponent *λ* of exponential function expressing daily new cases. As a result, the process from the start to the convergence of infection were clarified as the trends in isolation rate q and social distancing ratio x for each country. Through these analyses, the importance of dynamic parameters such as isolation rate q, increasing rate of cumulative cases(IRCC) and also increasing rate of cumulative PCR tests (IRCT), were emphasized in order to assess the dynamic transition of epidemic infection based on SIQR model.

- The increasing rate of cumulative cases (tested and isolated persons) IRCC was chosen and analyzed as a candidate for the isolation rate q with a practical and physical background,
- A normalization coefficient C was introduced in expression q=C-IRCC to normalize the isolation rate q, and the social distancing ratio x within the constraint of 0≤ q, x≤1. These constraints gave another constraint to normalization coefficient C with allowable narrow path between upper and lower boundaries. q and x were calculated for two types selected for C, a simple mode that traces the neutral line within this narrow path and a complex mode that traces the upper boundary curve in a similar fashion.
- Loci on the q-x plane in the simple mode shows a clear difference between countries. Taiwan and South Korea exhibit rapid increase in isolation rate in the early stages of infection, whereas countries in Europe exhibit the early increase in the social distancing ratio.
- In the complex mode in which the normalization coefficient C traces the upper boundary curve of the narrow path, all countries exhibit the linearly varying loci which is almost same with the result of the mathematical solution with the sequential calculation method simulating the bipedal walking in the previous report.
- The number of PCR tests was examined in relation with isolation rate, and the increasing rate of cumulative tests is found to be important as a means to promote the isolation rate.

## Data Availability

Only publicly available database was used.

https://ourworldindata.org/coronavirus-source-data

## Acknowledgment

The author would like to thank Professor Emeritus Ikuo Yoshihara of the University of Miyazaki for his detailed suggestions and discussions, and also Professor Emeritus Takashi Odagaki of Kyushu University for his SIQR model that inspired the author to complete these analyses.

